# Self-Supervised Contrastive Learning to Predict Alzheimer’s Disease Progression with 3D Amyloid-PET

**DOI:** 10.1101/2023.04.20.23288886

**Authors:** Min Gu Kwak, Yi Su, Kewei Chen, David Weidman, Teresa Wu, Fleming Lure, Jing Li

## Abstract

Early diagnosis of Alzheimer’s disease (AD) is an important task that facilitates the development of treatment and prevention strategies and may potentially improve patient outcomes. Neuroimaging has shown great promise, including the amyloid-PET which measures the accumulation of amyloid plaques in the brain – a hallmark of AD. It is desirable to train end-to-end deep learning models to predict the progression of AD for individuals at early stages based on 3D amyloid-PET. However, commonly used models are trained in a fully supervised learning manner and they are inevitably biased toward the given label information. To this end, we propose a self-supervised contrastive learning method to predict AD progression with 3D amyloid-PET. It uses unlabeled data to capture general representations underlying the images. As the downstream task is given as classification, unlike the general self-supervised learning problem that aims to generate task-agnostic representations, we also propose a loss function to utilize the label information in the pre-training. To demonstrate the performance of our method, we conducted experiments on the Alzheimer’s Disease Neuroimaging Initiative (ADNI) dataset. The results confirmed that the proposed method is capable of providing appropriate data representations, resulting in accurate classification.

## 1. Introduction

Alzheimer’s disease (AD), the most common form of dementia, is a degenerative and irreversible brain disorder. AD symptoms initially include a loss of short-term memory ability, and as the symptoms get worse, cognitive decline occurs. Many people are currently suffering from AD. It is estimated that there are 6.5 million individuals age 65 and older affected by AD in the United States alone. The number is also projected to reach 13.5 million by 2060 (Alzheimer’s Association, 2022). In addition, developing efficacious AD drugs is a challenging task with a high failure rate, and there is no disease-modifying treatment available other than the highly controversial Aduhelm. There is a strong consensus that the most effective treatment regime should target the early stages of the disease before irreversible brain damage has occurred (Cummings et al., 2020). Thus, the early identification of an individual’s condition is important.

Mild Cognitive Impairment (MCI) is a prodromal phase of the disease when individuals show some noticeable signs of memory and cognitive decline, but the symptoms are not severe enough to disrupt their daily activities. MCI is a high-risk stage that 10-15% of individuals progress to AD each year. It is important to identify which MCI individuals will progress/convert to AD within a short period of time (a.k.a. converters), which could be an important phase for early intervention to try to slow down the progression. This has been formulated as a classification problem (i.e., classifying MCI individuals into converters vs. non-converters) in AD literature.

Neuroimaging is a major tool for AD-related assessments. Among various neuroimaging modalities, volumetric magnetic resonance imaging (MRI) and positron emission tomography (PET) are widely used (Katabathula et al., 2021; Lu et al., 2018; Zhang et al., 2011). MRI can provide information about the structural alteration of the brain (Ritchie & Lovestone, 2002). PET can provide information about functional and pathological changes of the brain. A commonly used PET imaging modality is FDG-PET which measures cerebral glucose metabolism. Amyloid-PET has a favorable characteristic for AD diagnosis as it measures the accumulation of amyloid plaques in the brain – a hallmark of AD. It is of great interest to use amyloid-PET for converter vs. non-converter classification of MCI patients. A support vector machine was trained on fractal dimension and Shannon entropy as features extracted from amyloid-PET images for MCI conversion classification (Yan et al., 2019). Combining amyloid-PET and MRI with a multi-modality sparse representation-based classification method was proposed (Xu et al., 2016). A transfer learning method was proposed to exploit all the features of MRI, FDG-PET, and amyloid-PET data while being applicable to individuals with missing modalities (Liu et al., 2021).

There are two limitations of the existing work we want to tackle in this paper: First, the existing studies using amyloid-PET for MCI conversion classification focused on pre-defined features (e.g., regional amyloid measurements). Building a deep learning model that takes the 3D amyloid-PET images as input without feature engineering will greatly complement the existing studies. Second, it is a popular research area for building deep learning models based on neuroimages including MRI and PET in AD-related studies. However, most of the existing studies used fully supervised learning models that are trained using only labeled data, which do not explore other available data sources such as data without labels.

Self-supervised learning has gained popularity because of its superior capability of capturing general data representations from all available data sources. The latest achievements of self-supervised learning argue that the learned metric space implicitly captures the semantic similarities underlying the data, resulting in robust representations that are broadly transferable to various downstream tasks by fine-tuning. By learning the general representations through pre-training, a model that is less biased to the limited label information can be trained. It has resulted in great improvement in various domain applications, including but not limited to natural images (Caron et al., 2021), ultrasound video-speech (Jiao et al., 2020), histopathology images (Ciga et al., 2022), autonomous driving (Luo et al., 2021), and medical images (Chowdhury et al., 2021). Next, we introduce some recent studies using self-supervised learning in applications involved medical images. For example, the hybrid architecture of UNet and vision transformer, UNETR, was introduced to learn the sequence representations of 3D input for medical image segmentation. It achieved considerable performance gains for multi-organ, brain tumor, and spleen segmentation tasks (Hatamizadeh et al., 2022). Furthermore, UNETR was improved by adopting the Swin Transformer architecture for efficient training. It also introduced several tailored proxy tasks for proper self-supervised learning in the medical domain (Tang et al., 2022). There are a few self-supervised learning studies related to AD. Training a model with both functional MRI and structural MRI by contrastive fusion function was proposed for classifying AD and healthy controls (Fedorov et al., 2021). However, no study has been done to predict MCI conversion to AD using 3D amyloid-PET, which motivated our work in this paper.

In this study, we propose a self-supervised learning method to predict MCI conversion to AD with 3D amyloid-PET. We hypothesize that self-supervised learning to obtain general representations from all available data can help the model achieve better performance in the downstream classification task. Moreover, we introduce a loss function to provide more suitable representations for converter vs. non-converter classification by exploiting label information in the pre-training step. To demonstrate the effectiveness of the proposed method, we conducted experiments on the ADNI dataset and compared it with alternative methods.

The main contributions of this study can be summarized as follows:

- **Impact on the medical field**: We propose a self-supervised contrastive learning method for predicting MCI conversion to AD with the emerging 3D amyloid-PET images. To the best of our knowledge, this has not been studied in previous studies.
- **Methodological contribution**: We propose a contrastive loss function that allows a pre-trained model to provide data representations more suitable for the downstream classification. This is different from existing contrastive learning methods. We demonstrate the better performance of the proposed method compared to existing contrastive learning and supervised learning methods.

The remainder of this paper is organized as follows. In Section 2, we briefly review the base self-supervised contrastive learning model for our method. Then, we introduce the proposed loss function. Section 3 gives the experimental settings and results on ADNI data. Finally, Section 4 contains our concluding remarks and future research directions.

## 2. Proposed Method

We propose a self-supervised contrastive learning method to predict MCI conversion to AD with 3D amyloid-PET. The proposed method is based on the Momentum Contrast (MoCo) framework, which is a popular self-supervised learning method that has recently set a milestone with its great computational efficiency (X. Chen et al., 2020; He et al., 2020). MoCo basically aims to learn data representations that can be generally used in various types of downstream tasks without label information in the pre-training step. To further improve MoCO, we propose a contrastive loss function as the key component to learn the data representations more suitable for the downstream classification task by utilizing the label information in the pre-training step. In this section, we begin with a brief review of MoCo, and then introduce the proposed loss function which utilizes the given label information for converter vs non-converter classification.

Let 𝒟 = 𝒟_*L*_ ∪ 𝒟_*U*_ be a training dataset, where 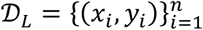 is the labeled data of *n* amyloid-PET images and 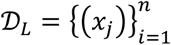 is the unlabeled data of *m* amyloid-PET images. A single-channel 3D amyloid-PET image is denoted as *x* ∈ ℝ^*d*×*h*×*w*^ where *d, h*, and *w* are the depth, height, and width of an image, respectively. In the pre-training step, MoCo trains a network with 𝒟 by discarding the label information to capture the general data representations. Then, the network is fine-tuned by 𝒟_*L*_ for a particular downstream task.

In the pre-training step, MoCo aims to learn the semantic data representations through instance discrimination without any label information. Namely, it assumes that each instance belongs to its own class. It is based on the intuitive concept of making similar instances pull each other together and dissimilar instances push each other away. Primarily, given an image *x*_*i*_ ∈ 𝒟, a stochastic data augmentation is applied to the same image twice to generate two different views: 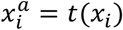 and 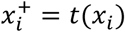, which are denoted as the anchor and a positive, respectively. These two views form a positive pair 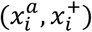 whose representations must be pulled together. Similarly, *K* negative examples 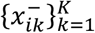 can be obtained by also applying *t*(⋅) to a subset of images sampled from 𝒟_*i*_ = 𝒟\{*i*}. By introducing a query network *f*_*θ*_(⋅) to achieve a nonlinear mapping *f*_*θ*_: 𝒳 → ℝ^*P*^ from the data space 𝒳 to a *P*-dimensional representation space, the contrastive loss function defined for the *i*^*th*^ image takes the following mathematical form:

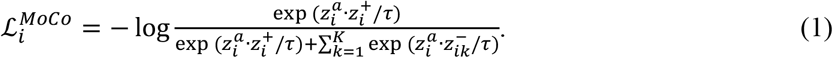

Herein, *τ* is a temperature hyperparameter for scaling, 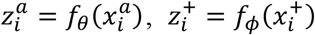, and 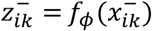, where *f*_*ϕ*_(⋅) is a key network. Note that *z* is L2-normalized, so the inner dot product *z* representations can be regarded as cosine similarity. We can intuitively understand that 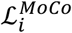 is related to the probability that the anchor is classified as the positive example with the same source image as itself among one positive and *K* negative examples.

The key network *f*_*ϕ*_ is structurally identical to *f*_*θ*_, but whose parameters *ϕ* are updated by an exponential moving average (EMA) of *θ*. Specifically, the update rule of key network parameters *ϕ* with a key momentum coefficient *λ* ∈ [0,1) is defined as follows:

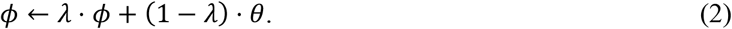

*λ* is typically set by a large value, such as 0.950 or 0.999, to slightly change the key representations. It maintains the consistencies for both positive and negative representations because they are generated from networks that are only different by a factor of 1 − *λ*. It consequently makes the instance discrimination task hard enough. It allows MoCo to generate proper and general data representations (He et al., 2020). Lastly, the core part of MoCo is the memory queue that stores past representations computed by the EMA-updated key network *f*_*ϕ*_. The memory queue enables MoCo to store and retrieve past representations easily during training with practicable computational costs. Note that storing *z* vectors mapped to a *P*-dimensional space in the memory queue makes MoCo computationally efficient.

By applying MoCo, we can additionally use the unlabeled MCI samples together with labeled MCI samples (i.e., samples with converter/non-converter information available). It is usually easier to obtain a large quantify of unlabled samples compared to labeled samples. It is also known that the classes are expected to be linearly separable on the pre-trained representation space if the samples are successfully discriminated and semantically well-clustered (Wu et al., 2018). Therefore, if desired, MoCo can be simply applied for converter vs. non-converter classification. However, we expect the model to have an additional characteristic more suitable for the classification task. To address this consideration, we propose a contrastive loss function that exploits the label information in the pre-training step.

The proposed loss function uses the negative examples in the memory queue with class label identical to the anchor as additional positive examples. Incorporating additional information as positive examples should be performed carefully. Selecting examples with different semantic representations from the anchor could easily break the overall learned representations. In our problem, the unlabeled data means the unknown state that has not yet been labeled. Therefore, additional positive examples could be obtained through methods such as pseudo labeling for unlabeled data, but we only aggregated the labeled data into positive examples. The proposed loss function is formulated as follows:

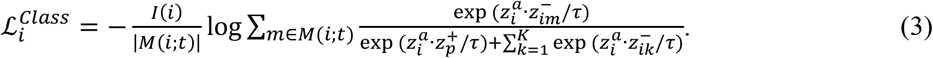

Herein, *I*(*i*) is an indicator function that *I*(*i*) = 1 if an anchor has a label, otherwise *I*(*i*) = 0. The proposed loss function can be applied when the anchor is labeled. *M*(*i*; *t*) refers to an index set of keys negative examples at iteration *t* having the same class as the anchor. Note that *M*(*i*; *t*) of the memory queue dynamically changes along the model training. By using the proposed loss function, samples with the same class label are expected to be similarly positioned in the representation space. Figure 1 demonstrates an overview of the proposed loss function.

**Figure 1.**
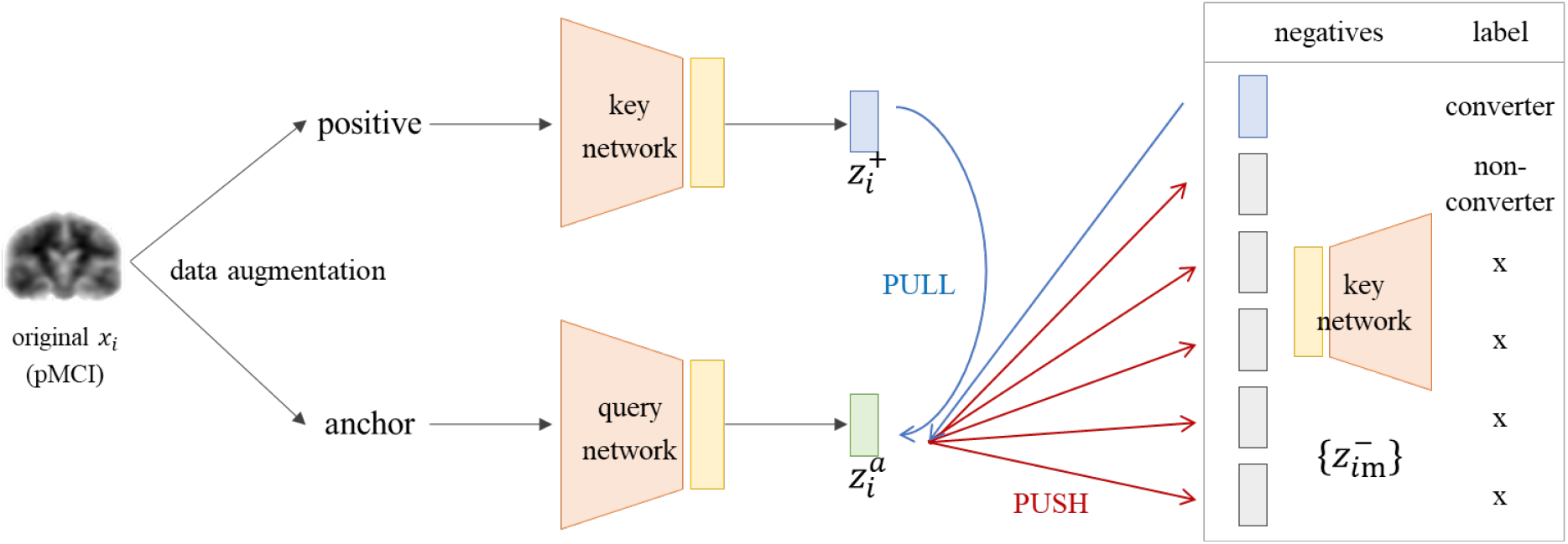
Graphical overview of the proposed loss function 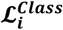. The labeled negative examples with the same class as the anchor are aggregated as additional positive examples.

The final contrastive loss function defined for the *i*^*th*^ image is formulated as follows:

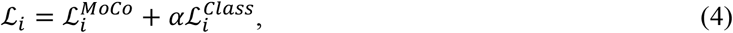

where *α* is a balancing hyperparameter. After the pre-training, the proposed model is fine-tuned on 𝒟_*L*_, the same as MoCo. Details are illustrated in Section 3.

## 3. Experiments

### 3.1. Data

To demonstrate the effectiveness and performance of the proposed method, we conducted experiments on the ADNI dataset. ADNI is one of the largest datasets for AD studies to date, with the primary goal being to test whether serial MRI, PET, other biological markers, and clinical and neuropsychological assessment can be combined to measure the progression of MCI and early AD. We downloaded 3D amyloid-PET imaging data from 612 individuals. The individuals who converted to AD within 36 months of MCI diagnosis were assigned as converters, and the individuals who did not convert were assigned as non-converters. There were 158 converters and 463 non-converters, and additionally 443 unlabeled MCI images. The unlabeled MCI refers to images that the AD conversion cannot be determined because there are no records at or after 36 months.

We also downloaded the T1-weighted MR image corresponding to each PET image from the ADNI database. The T1-weighted MR images were spatially normalized using the Computational Anatomy Toolbox 12 (CAT12) (Gaser et al., 2022) with Statistical Parametric Mapping (SPM12) (Ashburner et al., 2014) and a standard brain atlas from the Montreal Neuroimaging Institute (MNI). Then, each PET image was co-registred to the corresponding MRI. The spatially normalized PET images have a size of 121 × 121 × 121 and a voxel size of 1.5 mm in depth, height, and width. Then, we applied zero padding to generate 145 × 145 × 145 images to properly apply the data augmentation techniques commonly used in image analysis. Finally, we resized the images to a size of 72 × 72 × 72 using nearest-neighbor interpolation to reduce the computational cost.

### 3.2. Model Architecture and Training Hyperparameters

Based on recent works (Caron et al., 2020; T. Chen et al., 2020; He et al., 2020), a ResNet-50 encoder (He et al., 2016) with the fully connected layers replaced with a two-layer multilayer perceptron (MLP) was used for model architecture. The output dimension of *f* was set to 128. We used 3D ResNet-50, which increased one dimension of the convolutional filters of the existing architecture and replaced the first 7 × 7 × 7 convolution layer with a 3 × 3 × 3 convolution layer with a stride of one and zero padding of one.

We set the temperature *τ* = 0.2, the memory queue size *K* = 1024, and the key momentum coefficient *λ* = 0.950. Self-supervised pre-training was performed for 100 epochs with a batch size of 16. Using a base learning rate of 0.0001, the network was optimized using the AdamW (Loshchilov & Hutter, 2017) optimizer with a momentum of 0.9. The learning rate is gradually dropped to zero by following a half cosine schedule. MoCo originally uses a shuffling batch normalization technique to prevent the model from learning degenerate solutions by focusing on subtle cues induced by batch normalization. Because it requires multiple GPUs, we used ghost normalization (Hoffer et al., 2017). It divides each training batch into smaller sub-batches and normalizes them independently. We used ghost normalization with eight splits to mimic the behavior of MoCo trained on eight GPUs. For the proposed contrastive loss function, values in {0.25, 0.5, 1, 2, 3, 5} were used to investigate the effect of the balancing hyperparameter *α*.

For all experiments, we randomly split the labeled (converters and non-converters) data into 90% for training and 10% for testing while preserving the class distribution. The unlabeled data is used for additional training data only in the pre-training step. We organized the data so that the individual IDs in the training data are not included in the testing data. For all experiments, we conducted ten repeated trials with different random seeds and reported the average values of the corresponding evaluation metrics.

### 3.3. Representation Quality Evaluation

We first conducted an experiment comparing the performance of the vanilla MoCo and the performance of the proposed model according to the hyperparameter *α* in Eq.(4). Investigating the effect of the balancing hyperparameter used for introducing a new loss term is an essential task because it strongly affects the model performance in general. We evaluated the representation quality by *k*-nearest neighbor classification (*k*-NN), a widely used evaluation protocol (Wu et al., 2018). We froze the pre-trained model to achieve the representations of the training data for *k*-NN. Then, the *k*-NN classifier matches the label of a testing sample to the predicted label from *k*-nearest training images. We used the area under the receiver operating characteristics (AUROC) to appropriately evaluate the models with class imbalance. AUROC is a threshold-independent metric that measures the trade-off between sensitivity and specificity at all thresholds. Table 1 shows the *k*-NN results over different values of *α*. The performance of MoCo that uses only ℒ^*MoCo*^ is also reported as a baseline. We used *k* of five.

**Table 1.** 5-NN classification performance obtained from MoCo and the proposed method over different values of *α*. The best result is **boldfaced**.

| Model | MoCo | Proposed Method |  |  |  |  |  |
| --- | --- | --- | --- | --- | --- | --- | --- |
| $\alpha$ | 0 | 0.25 | 0.5 | 1 | 2 | 3 | 5 |
| AUROC | 76.88 | 79.49 | 80.41 | **80.99** | 80.71 | 79.26 | 79.24 |

As shown in Table 1, it can be observed that the proposed loss function enhances the representation quality over MoCo regardless of the value of *α*. The performance of the proposed model is about 2.5% higher than that of MoCo, even when the lowest (*α* = 5). We can confirm that the proposed loss function helps the model to provide more appropriate representations regardless of the balancing hyperparameter because it is designed for performing the given classification task well. The best representation is obtained when *α* = 1. Compared to MoCo, the best 5-NN result is about 4% higher. Furthermore, it can be observed that the model performance gradually decreases when *α* is greater than one. Our interpretation is that the model focused too much on aggregating the instances of the same class so that the instance discrimination task was not properly done. The model was not able to capture the general characteristics of data because of the trade-off. Based on the result, we fixed *α* in the remaining experiments.

Moreover, we compared the representations of MoCo and the proposed model by visualization. We visualized the high-dimensional representations by reducing them to two-dimensional vectors by uniform manifold approximation and projection (UMAP) (McInnes et al., 2018). Figure 2 shows the UMAP results. Grey, blue, and red points refer to the unlabeled, converter, and non-converter training data, respectively. As shown in Figure 2 (a), converter and non-converter samples are overlapped more when MoCo was simply used. During learning the semantic structure of the data, converters and non-converters were separated to some extent because they have different amyloid-PET characteristics. However, because MoCo is designed to produce general representations, not for specific classification task, the classes were not separated enough. Compared with Figure 2 (b), we can confirm that the proposed model more clearly separated converters and non-converters. Most of the converter points are located close to the edge of the cluster. Thus, better performance can be expected if the classifier is trained with the representations.

**Figure 2.**
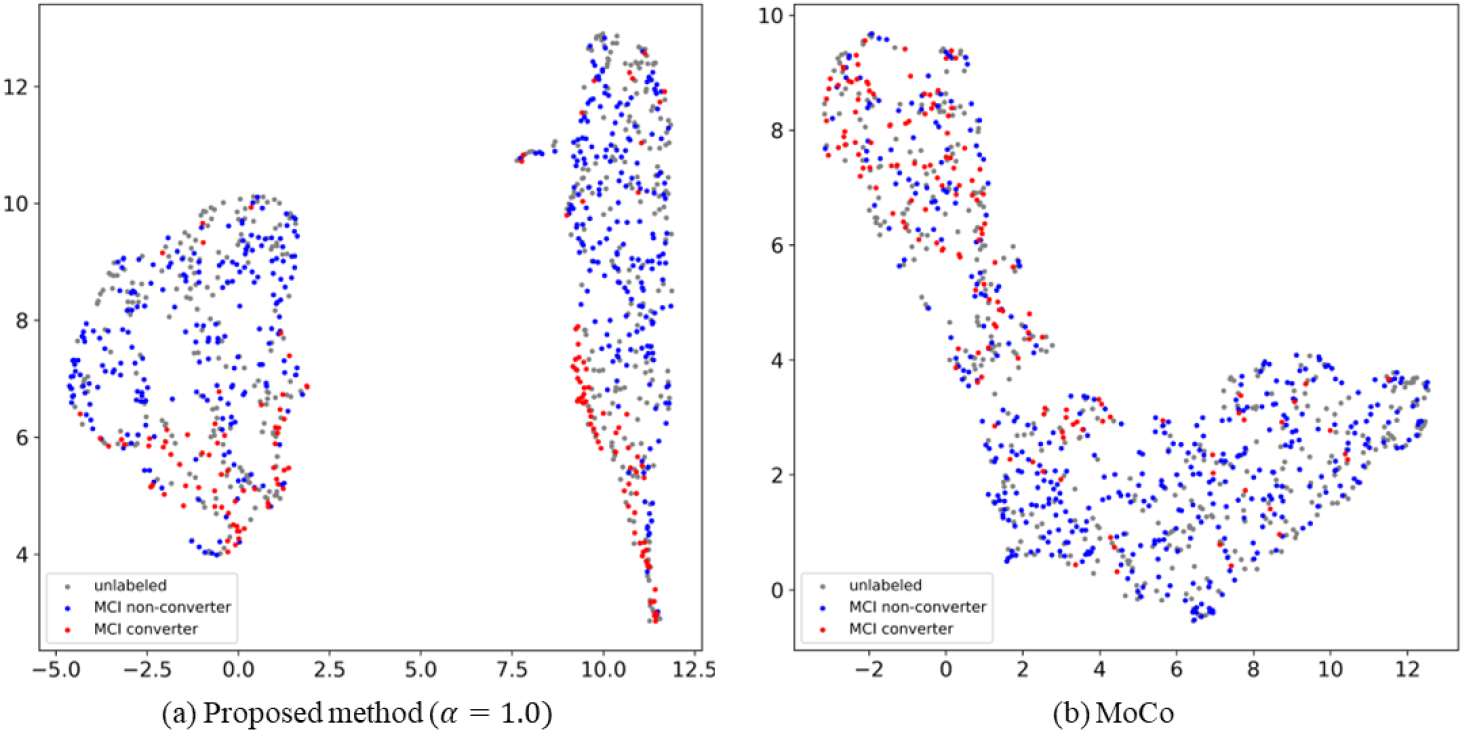
UMAP visualization of pre-trained representations of training data. (a) The proposed model; (b) Vanilla MoCo. Grey, blue, and red points refer to the unlabeled, converter and non-converter data, respectively.

### 3.4. Clasification performance and comparison

Finally, we compared the classification accuracy of the proposed method with supervised learning and MoCo. Supervised learning refers to the conventional model that is trained with only labeled data 𝒟_*L*_. It is trained for 100 epochs with an AdamW optimizer using an initial learning rate of 0.0001. The learning rate is also decayed to zero using a half cosine schedule. The batch size was set to 16. Because our amyloid-PET data has class imbalance, supervised learning tends to be overfitted to the majority class. We applied a resampling technique in every mini-batch to address the class imbalance problem. We also used an adaptive cutoff strategy to fairly select the threshold for classification probability under the class imbalance. Specifically, we created the ROC curve and selected the threshold from the point closest to the corner point with a false positive rate of zero and a true positive rate of one.

To evaluate the classification performance of MoCo and the proposed method, we conducted a supervised fine-tuning to train a classifier. The MLP layer was replaced with a single-layer softmax classifier. Unlike the *k*-NN evaluation protocol, the pre-trained network was not frozen. We fine-tuned the network on 𝒟_*L*_ for 10 epochs. Except for the number of epochs, the remaining hyperparameters for fine-tuning were used the same as those of supervised learning. In addition, we trained a random forest classifier (Breiman, 2001) with the generated representations from the frozen pre-trained proposed model. Training a machine learning classifier based on a self-supervised learning model is commonly used (Purushwalkam & Gupta, 2020). Because tuning hyperparameters for the pre-training step requires considerable time, it is proper to use a relatively simple model. Thus, we selected the random forest. We used evaluation metrics – AUROC, accuracy, sensitivity, and specificity – all widely used for medical image analysis.

Table 2 presents the classification results. Overall, classification performance is good in the order of the proposed model, MoCo, and supervised learning. It can be noticed that simply using MoCo enhanced the model performance in terms of AUROC and sensitivity. It improved the prediction of the minority class, converters. We can confirm that the general data representations learned from self-supervised learning lead to classification performance improvements.

**Table 2.** Classification performance. The best value for each metric is **boldfaced**.

| Method | AUROC | Accuracy | Sensitivity | Specificity |
| --- | --- | --- | --- | --- |
| Supervised | 0.8166 | 0.7777 | 0.7324 | 0.7901 |
| MoCo & Finetuning | 0.8268 | 0.7714 | 0.7403 | 0.7812 |
| Proposed & Random Forest | 0.8511 | 0.7945 | 0.7529 | 0.8062 |
| Proposed & Fine-Tuning | **0.8528** | **0.8119** | **0.7743** | **0.8228** |

Moreover, the proposed contrastive loss function further improved the performance of MoCo. Both training a random forest classifier and applying fine-tuning showed better performance than comparable models. Fine-tuning achieved the best performance with considerable gains of about 2.5% of AUROC, 4% of accuracy, 3.5% of sensitivity, and 4% of specificity than MoCo. This in turn proves our original conjecture that exploting the information of negative samples with the same class provides useful representations appropriate to the downstream converter vs. non-converter classification. The combination of general representations by instance discrimination and classification-specific representations by the proposed contrastive loss function brought a substantial performance gap.

## 4. Conclusions

In this paper, we proposed a self-supervised contrastive learning method for predicting MCI conversion to AD with 3D amyloid-PET images. Amyloid-PET images have favorable characteristics for early AD diagnosis, which is critical in the dementia research field. We used 3D PET images to avoid using feature engineering that requires domain knowledge and related tools. Applying self-supervised learning prevents the classification model from being biased on the limited given label information. Moreover, we proposed a self-supervised contrastive loss function to generate suitable data representations for the downstream classification task of converters and non-converters. The proposed loss function utilizes the negative labeled examples of the same class as the anchor for additional positive examples. The experimental results showed that the proposed method contributes to considerable classification performance gains. To the best of our knowledge, this is the first study to apply self-supervised learning for predicting MCI conversion to AD with 3D amyloid-PET images.

There are several intriguing directions to extend our work. First, we can jointly use the demographic information for pre-training. Many existing studies on MRI and PET use clinical information to further improve model performance. As self-supervised learning research on tabular data has been actively conducted, better data representations can be obtained using clinical information. It can also be used as a critical cue to select additional positive examples. Second, we can develop a multi-modal contrastive learning framework for both MRI and PET. We can expect the model learns the brain structure characteristics from MRI modality and the accumulation level of amyloid plaques from PET.

## Data Availability

All data produced are available online at ADNI (https://adni.loni.usc.edu).

## Acknowledgments

This research is supported by NIH grant 2R42AG053149-02A1 and NSF grant DMS-2053170.

## Notes

### Competing Interest Statement

The authors have declared no competing interest.

### Author Declarations

The study used (or will use) ONLY openly available human data that were originally located at Alzheimer's Disease Neuroimaging Initiative (ADNI).

